# Factors influencing the implementation of integrated screening for HIV, syphilis, and hepatitis B for pregnant women in Nepal: a qualitative study

**DOI:** 10.1101/2024.02.26.24303120

**Authors:** Lucie Sabin, Hassan Haghparast-Bidgoli, Bibhu Thapaliya, Obindra Chand, Sanju Bhattarai, Abriti Arjyal, Naomi Saville

## Abstract

According to the Nepalese national guidelines, integrated screening for HIV, syphilis and hepatitis B should be offered free of charge to all pregnant women during their first antenatal visit. However, the screening uptake among pregnant women remains low in the country. Identifying factors influencing the implementation of integrated screening for HIV, syphilis, and hepatitis B is essential to increase uptake and prevent mother-to-child transmission. This study investigated the knowledge, attitude, and perceptions of pregnant women, their family members, healthcare workers and decision-makers on integrated antenatal screening. On the demand side, we conducted 26 semi-structured in-depth interviews with pregnant women, their husbands, and mothers-in-law in Kapilvastu and Kathmandu. On the supply side, we conducted 11 interviews with health workers involved in antenatal screening and local and national health system decision-makers. Data were analysed using a thematic content analysis. A combination of the social-ecological model and the WHO building blocks provided a theoretical framework for interpreting the data. The analysis showed that integrated antenatal screening for HIV, syphilis and hepatitis B in Nepal involved many stakeholders and was influenced by various factors. Barriers were mainly on the supply side, including a lack of resources, a shortage of healthcare workers and a lack of training. Husbands and in-laws also play an important role in the acceptance of screening by pregnant women, mainly in rural areas. Stigma and discrimination against people with sexually transmitted diseases were reported high in the communities, and knowledge of hepatitis B and syphilis was low. Improving integrated antenatal screening will require a multi-sectoral approach with greater engagement with communities through awareness programs and enhancement of the role of health posts. This study may be useful to inform decision-makers about the challenges and enable affecting integrated screening, to guide the design of targeted interventions to improve antenatal screening rates.

## Introduction

Human Immunodeficiency Virus (HIV), syphilis and hepatitis B are major contributors to the global burden of disease and disability. In 2017, 5.2 million people were living with HIV in the Asia Pacific region (1). In 2012, an estimated 1.8 million women were infected with syphilis in the Southeast Asia region, including Nepal (2) and 39 million people with hepatitis B with a prevalence of 2.0% (3).

Information on the prevalence of these sexually transmitted diseases (STDs) in Nepal are scarce. In 2020, HIV prevalence is estimated at 0.12% among adults aged 15-49 years (4). In 2019, the prevalence of hepatitis B was 0.9% (5). The majority of official data available for syphilis and hepatitis B are for specific population groups (e.g. sex workers and drug users) and no data available at national level (4).

HIV, syphilis and hepatitis B can be transmitted from infected mothers to their children during pregnancy, childbirth or breastfeeding also known as mother-to-child transmission (MTCT) or vertical transmission. An estimated 10,000 new HIV infections occurred among children aged 0–14 years in the Asia Pacific region in 2017 (1) and 1.3 million pregnant women are at risk of transmitting hepatitis B to their newborns each year (3). The global number of adverse pregnancy events attributable to maternal syphilis infection was estimated to 52,307 cases in the Southeast Asia Region and 13,472 in the Western Pacific Region (6). Screening during antenatal care (ANC) is a key to eliminating MTCT. Early detection of these diseases in pregnant women and appropriate treatment can drastically reduce the risk of vertical transmission (7–9). Since 2010, an estimated 7,400 new HIV infections among children in the Asia Pacific region were averted because of interventions aimed at reducing the MTCT of HIV (1).

To guide a path towards the triple elimination of MTCT of HIV, syphilis and hepatitis B in Asia and the Pacific, the World Health Organisation (WHO) developed a regional framework (10). This framework aims to eliminate these three infections in newborns and infants by 2030 in Asia, emphasising the principle of care centred on the mother, newborn and child, and universal health coverage. The framework encourages an integrated approach to triple elimination, recognising the interconnectedness of the three diseases and the potential for resource optimisation. Integrated antenatal screening for HIV, syphilis and hepatitis B allows pregnant women to be tested for these three diseases with a single blood sample.

In Nepal, screening for HIV during pregnancy is included in the National HIV Testing and Treatment Guidelines and it is mandated to be offered free of charge to all pregnant women during their ANC visits (11). The guideline also states that all adults should be offered STD services while receiving HIV services. In addition, according to the 2030 roadmap for safe motherhood and newborn health in Nepal (12), HIV and syphilis screening should be routinely carried out together during ANC visits for pregnant women. However, screening for HIV remains low in Nepal. Out of a total of 755,647 pregnant women in 2019, 431,912 were tested for HIV (4). Limited information is available on the screening of syphilis and hepatitis B. On supply side, only 16 percent of facilities have the capacity to screen for syphilis infection (11) but no data is available for hepatitis B. Yet unknowingly infected women may transmit infections to their sexual partners or children through MTCT. This also prevents them from accessing timely treatment leading to long-term complications that generate significant costs to the health system. In addition, low uptake of STDs screening services can exacerbate existing health disparities, with vulnerable populations, such as marginalised communities or migrant populations, facing additional barriers to accessing screening services.

Many stakeholders are involved in the screening of HIV, syphilis and hepatitis B of pregnant women including healthcare workers on the supply-side and pregnant women’s family members on the demand-side. Thus, as well as investigating the knowledge, attitude, and perceptions of pregnant women on integrated antenatal screening, it is essential to consider those of their family members and healthcare workers. Few studies in Nepal investigated demand side (13,14) and supply side (15) affecting antenatal screening of HIV. Roshna Thapa et al. (2019) (14) examined the association between women’s empowerment and HIV testing among Nepali women using data from the 2012 Nepal Demographic and Health Survey. They found that age, education, and wealth were factors that determined HIV testing among women. Pokharel, Abbas, et Ghimire (2011) (13) studied the response to the implementation of the prevention of mother-to-child transmission programme and found that the acceptability of the test after counselling was satisfactory. They stressed the need for greater male involvement. Only one study examined supply-side barriers. Using the 2015 Nepal Health Facility Survey (NHFS), Acharya et al. (2020) (15) showed that the readiness of facilities for HIV counselling and testing services is higher than for STD services. They found persistent gaps in staffing, guidelines, drugs, and commodities in both services.

To the best of our knowledge, no study conducted in Nepal has investigated factors influencing integrated antenatal screening of HIV, syphilis, and hepatitis B. By interviewing pregnant women, their family members, health workers and decision-makers, this study aimed to explore the barriers and enabling factors to integrated antenatal screening for HIV, syphilis, and hepatitis B in Nepal. This study may be useful to inform decision-makers about the challenges and facilitators of integrated screening, guide the design of targeted interventions to improve antenatal screening rates in the country by removing potential barriers, and contribute to the global effort to improve maternal and child health by promoting integrated antenatal screening for HIV, syphilis, and hepatitis B.

## Methods

### Setting

Nepal’s health budget has risen from NRs 23.81 billion in 2011 to NRs 122.79 billion in 2022, and the share of health expenditure in consolidated public spending has reached 7.5% of GDP, or 30% of current health expenditure. In 2021, external healthcare expenditure and out-of-pocket expenditure remained important, representing respectively 10% and 54% of current healthcare expenditure.

Many health workers from different health facilities are involved in the provision of screening for STDs for pregnant women in Nepal. Female Community Health Volunteers (FCHVs) play an important role in antenatal screening in rural communities. They link communities and the health system, providing basic health services and health awareness training. Health posts are the most available type of institution for primary healthcare in the communities across Nepal. Women go there for registration, ANC visits, screening, and immunisation as part of their pregnancy. Public hospitals have a larger capacity than health posts and often have more screening facilities. They are owned and operated by the government, while private hospitals are owned and operated by private entities. Other essential actors in antenatal screening in Nepal are non-governmental organisations (NGOs). Some are involved in screening at-risk individuals, treating, and monitoring positive cases.

For the purposes of our study, it is important to understand the differences between the Kathmandu Valley and Kapilvastu district where our study took place.

The Kathmandu Valley, with Kathmandu the federal capital city of Nepal, is the most developed urban area of Nepal with about more than 1.5 million population (16). Located in Bagmati province in the hills, the agglomeration attracts migrants from all over the country. The Kathmandu Valley houses many of the health facilities in the country with 30 public hospitals including the Paropakar Maternity and Women’s Hospital (PMWH) - the national referral centre for obstetrics and numerous other tertiary referral hospitals and 82 health posts (17).

Kapilvastu is a *Terai* (plains) district, on the border with India, which has a 55-bed government district hospital and 12 health posts. Its geographical position makes it a district with a large population of Muslims, Madhesi Dalits and Hindu Indian origin ethnic groups. These groups are often marginalised and face discrimination (18). Male foreign labour migration to India, the Middle East and East Asian countries (especially Malaysia) (19) is widespread in this district which increases vulnerability to STDs. Most of the population are Awadhi-speaking rural farmers (20). The Terai area of Nepal has a low Human Development Index (HDI) score largely driven by poor education indicators (21) and the highest levels of gender disparity in Nepal (18).

### Participants recruitment

Characteristics of participants are presented in Table 1. Participants were recruited between the 10 January 2023 and the 7 April 2023. On the demand side, we conducted a total of 14 semi-structured in-depth interviews with main users or beneficiaries (i.e. pregnant women) and their household members (i.e. their husbands and mothers-in-law) in Kapilvastu and 12 in Kathmandu. We interviewed 12 currently pregnant women (6 in Kapilvastu and 6 in Kathmandu) and since joint households is widespread in Kapilvastu, we also interviewed 4 mothers-in-law. To explore male spouses’ perspectives on maternal health and antenatal screening for STDs we interviewed 10 husbands (4 in Kapilvastu and 6 in Kathmandu), among them, five were husbands of pregnant women interviewed.

**Table 1.** Participants and required number of Semi-Structured Interviews (SSI)

|  | Activity | Criteria of selection | Kathmandu | Kapilvastu |
| --- | --- | --- | --- | --- |
| Demand side |  |  |  |  |
| Pregnant women | 12 SSIs | Woman from disadvantaged caste. | 3 | 3 |
|  |  | Woman from high caste. | 3 | 3 |
| Husbands | 10 SSIs | Husband from disadvantaged caste. | 3 | 2 |
|  |  | Husband from high caste. | 3 | 2 |
| Mothers-in-law | 4 SSIs | Mother-in-law from disadvantaged caste. | - | 2 |
|  |  | Mother-in-law from high caste. | - | 2 |
| Total |  |  | 12 | 14 |
| Supply side |  |  |  |  |
| Health workers | 7 SSIs | Health worker from public hospital | 1 | 1 |
|  |  | Health worker from a stand-alone HIV testing site. | 1 | 1 |
|  |  | Health worker from a health post. | 1 | 1 |
|  |  | Health worker from a private hospital. | 1 | - |
| Decision-makers | 4 SSIs | Rep' from the Ministry of Health. | 1 | - |
|  |  | Rep' from the health division of the Municipality. | - | 1 |
|  |  | Rep' from the Nepal Health Sector Support Program. | 1 | - |
|  |  | A person implementing programs for the Global Fund. | 1 | - |
| Total |  |  | 7 | 4 |

In Kapilvastu, we contacted pregnant women through the Female Community Health Volunteers (FCHVs) purposively approached in one semi-urban municipality (Kapilvastu) and two rural municipalities (Shuddhodhan and Maya Devi). A trained and experienced qualitative researcher (BT) worked with five females and a male local interviewer to locate and interview pregnant women and their family members. They interviewed people of the same gender respectively. We trained local interviewers based on our review of the literature on how to conduct interviews on sensitive topics (22,23). We conducted nine of the interviews in the local language Awadhi and five in Nepali. A previous study in a similar context overcame the reluctance of mothers-in-law to let their daughters-in-law be interviewed in private by implementing a parallel interview strategy (24). We applied a similar strategy by conducting concurrent but separate interviews in two different locations of the household with pregnant women and their mothers-in-law. We interviewed all the husbands alone around the household.

In Kathmandu, we identified and approached participants during their visits to the ANC clinics of the PWMH. A gynaecologist from the hospital introduced us and the study to the participants in the ANC unit. Pregnant women and husbands were interviewed in a café nearby where we could ensure privacy respectively by BT and a male qualitative researcher trained for this study. Participants gave their written consent to participate. Pregnant women and husbands were relatively reluctant to participate in the study as we met them in the hospital setting, they were often rushing to go home after their check-up. 2 pregnant women and 4 husbands refused consent. We audio recorded all interviews which lasted 30 minutes to 2 hours.

On the supply side, we interviewed health workers involved in antenatal screening and local and national health system decision-makers. We conducted 4 interviews in Kapilvastu and 7 in Kathmandu. Participant characteristics are presented in Tables 3 and 4.

**Table 3.** Health workers’ characteristics.

| Nº | Organisation | Municipality | Occupation | Years of experience | Gender |
| --- | --- | --- | --- | --- | --- |
| 1 | Health post | Kapilvastu | Auxiliary Nurse Midwife | 6 | Female |
| 2 | NGO | Kapilvastu | Counsellor at ART Centre | 5 | Female |
| 3 | Public hospital | Kapilvastu | Nursing Officer | 7 | Female |
| 4 | Health post | Kathmandu | Auxiliary Nurse Midwife | 23 | Female |
| 5 | Public hospital | Kathmandu | Doctor | 7 | Male |
| 6 | Public hospital | Kathmandu | Auxiliary Nurse Midwife | 38 | Female |
| 7 | Private hospital | Kathmandu | Registrar in Obstetrics and Gynaecology | 3 | Female |

**Table 4.**
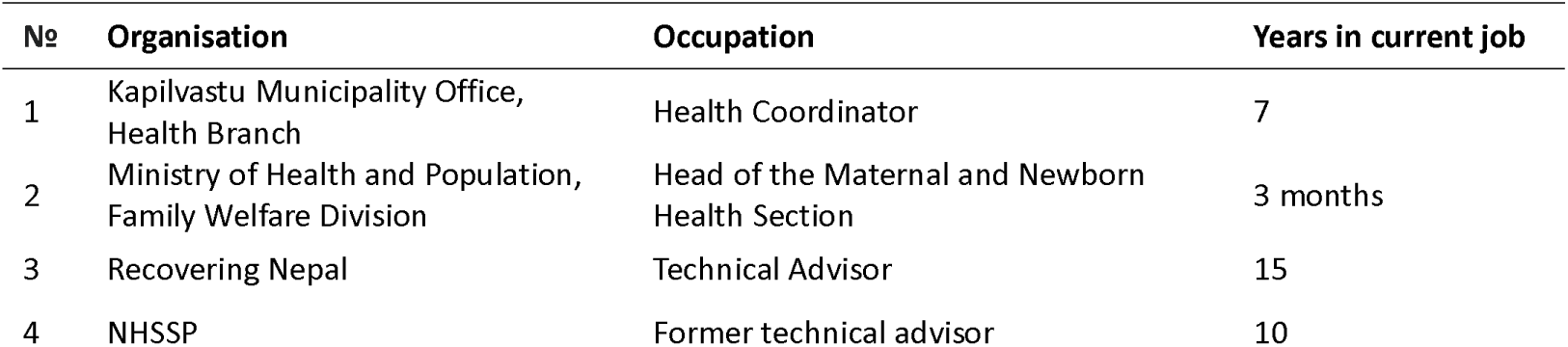
Decision makers’ characteristics.

We interviewed any consenting mothers-in-law and husbands of a sampled pregnant woman. In Kapilvastu, the strategy of interviewing pregnant women, husbands and mothers-in-law from the same family was prioritized to allow triangulation resulting in a household with all three household members interviewed, three with mother-in-law and pregnant women only interviewed and three with husband and pregnant women interviewed. Since not all women’s husbands and mothers-in-law were available at the time of interviews, husbands and mothers-in-law from different families were contacted until the sample size was met. This strategy of interviewing people from the same household was not appropriate in the Kathmandu urban context as pregnant women were reluctant to talk knowing that their partner would also be interviewed. Although pregnant women were usually accompanied by their husbands, the latter were in a hurry to get back to work. We were unable to interview mothers-in-law because they did not accompany pregnant women on ANC visits. Saturation was reached by the end of the data collection and new analytical insights from additional interviews became rare.

Pregnant women and their families were interviewed about their experience of ANC visits, blood tests, knowledge of and behaviours toward STD and antenatal screening for STDs during pregnancy. Our topic guides evolved after the pilot phase and over time, to explore emergent themes over the course of data collection. Vignettes, which allow for less personal exploration of sensitive topics that participants might otherwise find difficult to discuss (25–27), were used to interview pregnant women and mothers-in-law. In the vignette, participants were presented with a hypothetical situation in which they were discussing ANC visits and blood tests with a friend. In this hypothetical situation, the friend was also asking for advice about a genital discharge and the possibility of a STD. They aimed to reveal people’s beliefs and potential behaviours without being too intrusive. The vignette was designed according to the local context and on the advice of local researchers and is presented in S1 File.

### Data analysis

Data were analysed by LS and BT using NVivo to undertake thematic content analysis (28). A combination of the social ecological model and the WHO building blocks provided a theoretical framework for interpreting the data (Fig 1). The social ecological model proposes a holistic approach and considers the relationship between individuals and their environment to understand health-related behaviours (29). The model acknowledges that an individual’s behaviour is shaped through complex and interrelated level factors. In this study and according to McLeroy et al. (1988) (29), five levels that influence behaviour will be considered. The first level of influence is the individual-level. This level refers to the personal characteristics of an individual that influence their behaviour such as age, gender, ethnicity, knowledge, attitudes, skills, and beliefs. The second is interpersonal level, which describes the relationships between individuals and the social networks they belong to, such as family, friends, and neighbours. The third is institutional level that includes formal and informal structures of health in this particular study. The fourth is community level, which refers to established norms and values, standards, and social networks. The higher level of influence is the national level, describes how the laws, regulations, and policies affect behaviour by creating or limiting opportunities and resources, shaping social norms, and changing the physical and social environment. The WHO building blocks approach (30) was also used to analyse the data from the supply side perspective and was integrated to the social ecological framework. This approach provides a framework to improve the performances of health systems by considering six constituent elements: service delivery, health workforce, health information, medical products, health financing, and leadership and governance.

**Figure 1.**
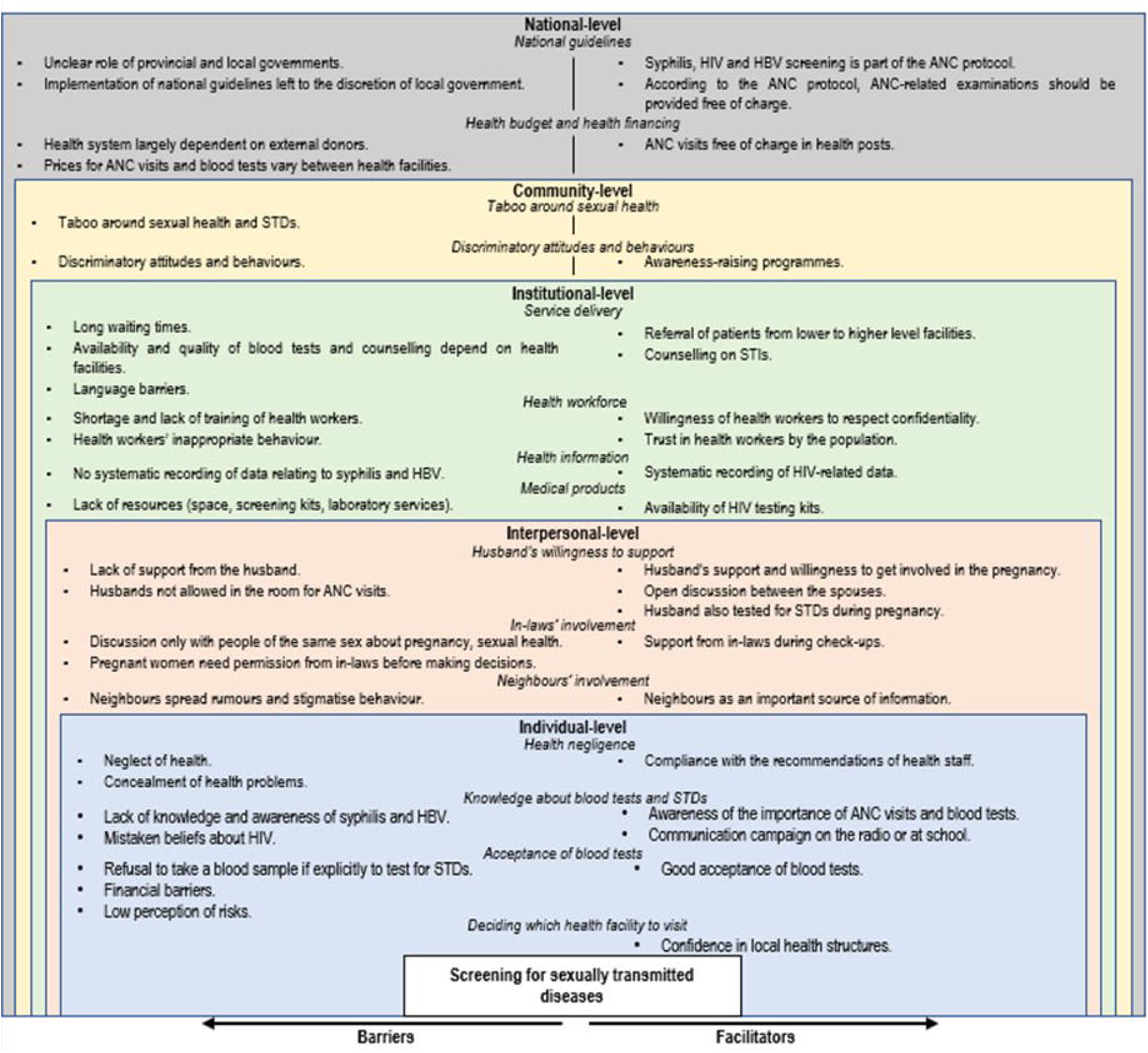
Combination of the social ecological model and the WHO building blocks.

After a step of familiarisation with the data, we read transcripts and carried out a line-by-line analysis by applying the different categories of the social ecological framework and the WHO building blocks in NVivo. To limit cultural bias and ensure rigor, we double coded some of the transcripts before finalising the main thematic framework and coding transcripts in NVivo. To develop a deeper analysis of the data, we also conducted a framework analysis (31). The codes identified in the thematic analysis were applied to the whole data in a systematic way. The themes were compared across the transcripts and specifically the different groups, to establish the range and similarities of the participants’ perceptions, experiences and views. We tabulated quotes from the transcripts within and between transcripts to triangulate the data. We reported results in accordance with qualitative research guidelines (32,33) and the COREQ checklist (32).

This study has been conducted under an extension of the UCL Global Engagement funding for the Comprehensive Anaemia Programme of Preventive Therapies (CAPPT) project. We received ethical approval for this study from the Nepal Health Research Council and University College London Ethics Committee.

## Results

We present the perspectives of the different participants together by comparing similarities and differences according to their localities and their characteristics. Results were presented according to the categories of the social ecological model (29) and the WHO building blocks (30). Fig 2 summarises the findings according to the social ecological model and the WHO building blocks.

**Figure 2.**
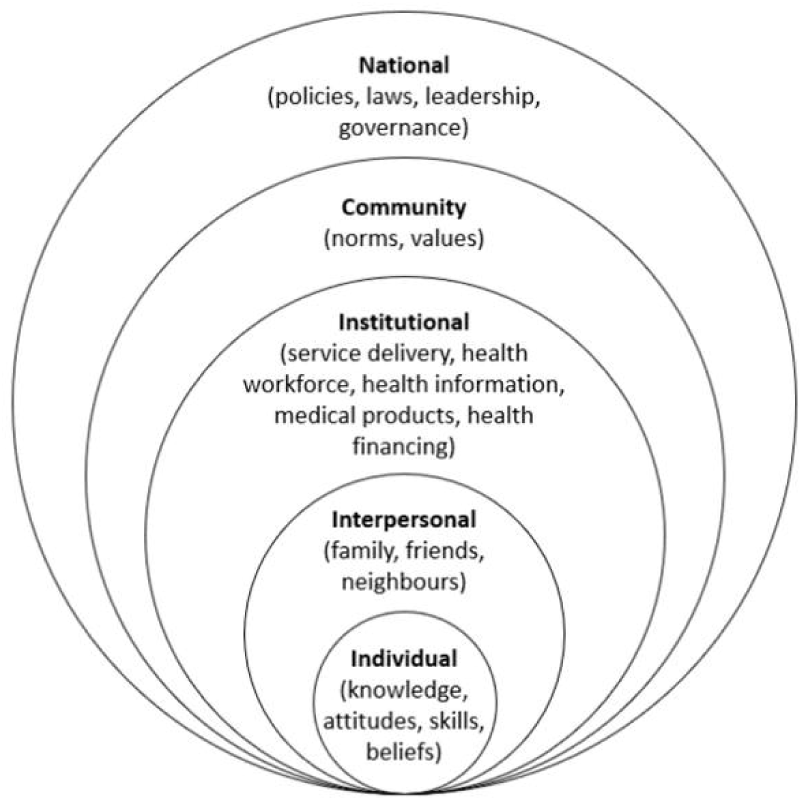
Factors influencing access to and provision of antenatal screening for HIV, syphilis and hepatitis B in Nepal presented according to the social ecological model and the WHO building blocks.

### National level

The WHO building block (30) on leadership and governance considers the policies, regulations and institutions that oversee and guide the health system and ensure the efficient and equitable provision of health services.

#### National guidelines

Syphilis, HIV, and hepatitis B screening is part of the ANC protocol in Nepal. However, in practice, STDs screening is not available in all facilities, in particular, rural health posts. This is despite claims to the contrary by the decision-maker interviewed from the Family Welfare Division of the Ministry of Health. *[PM2: HIV screening is conducted as a routine practice in Nepal, I remember it being implemented for a very long time, for more than 20 years, I think. Hepatitis B and Syphilis are also conducted routinely in every health institution in Nepal.]* The implementation of elements included in the national guidelines is left to the discretion of the local government. They have to convince their elected representatives to give them a budget for it. The services included in ANC package and the charges are decided by hospital management committees and by the municipality in the case of health posts. In the context of Nepal’s federalization introduced in 2015, the role of provincial and local governments is still unclear. *[PM3: For a long time, it was not clear whether the central or the federal government should look over the health office of the districts. Finally, the Sthaniya Ain [Local Law] has been introduced. It clarifies the services that fall under the local government. However, it’s still confusing whose responsibility it is to procure materials for the provincial level.]*

### Health budget and health financing

The financing building block (WHO 2007) concerns with raising adequate fund for healthcare system, allocation of resources, and protect people from high costs of care seeking.

The Nepalese health system is largely dependent on external donors. *[PM3: The revenue collected by the Government of Nepal is not sufficient to run all the health programs. From what I know, the Government of Nepal has enough money for regular salaries, management and running some programs but the rest of the money is given by donors.]* Based on consultation with its various bodies, diplomatic and policy sectors, the Ministry of Health and Population (MoHP) decides on health priorities. It discusses at the national level how health projects can be financed and whether it will be necessary to seek external funding to meet the needs.

The ANC protocol also states that any ANC-related examination should be provided free of charge. However, STDs screening is not always available for free. *[HW7: When I was working in the maternity ward, there was a minimum charge. It was lower than in other hospitals, but it was not free.]* At the level of health facilities, municipalities make decisions for health posts regarding the budget allocated, the purchase of equipment such as test kits and the services to be provided free of charge. *[PM4: If it is a protocol guideline which is not mandated by the Constitution, then it is the discretion of the local government or the health coordinator to implement it. First, they need to be convinced that it is important. Then, they need to convince their elected members to give them a budget for implementation. They can ignore it and not implement it because they can choose to do something much more appealing. Building a facility might be more appealing than just adding a small service which no one really knows.]* Public hospitals are funded by provincial governments. The management committee of each hospital has the power to decide and prioritise which services to include in the ANC package and which services to provide free of charge, based on the recommendations in the government protocols. It must coordinate with municipal public health officials and doctors to make a service mandatory.

ANC visits are free of charge at health posts. In public hospitals, the price varies from one institution to another and depends on the content of the examinations. *[HW7: Costs in public hospitals are low but they depend on the programs that the government is running.]* Participants complained about the price of ANC visits in private hospitals. *[PW4: In private health facilities, they take a lot of money, so it is better to go to health posts or government hospitals.]* This difference in cost justifies participants’ preference for public health facilities over private ones. *[PW8: I did everything in a private hospital. I only came here [PMWH] three months ago because of monetary issues. It was too costly in the private hospital.]*

When available at health posts, blood tests are provided free of charge. As with ANC visits, the price of blood tests varies from one public hospital to another. Blood tests are more expensive in private facilities, and patients complain about this to doctors. *[HW7: In private hospitals, serology for hepatitis B is considered to be very expensive in comparison to HIV because it requires more equipment. Patients come and tell us: “these three tests are expensive, doctor”.]* It is common for patients to go to private facilities for ANC visits, but to go to public facilities for blood tests because they are cheaper. *[PW8: I was advised by the doctor that it’s costly in the private hospital, so I went to a laboratory in Baneshwor.]* Participants cited money as a reason for refusing blood screening during pregnancy. *[PW6: I have not done blood tests because I don’t have money. If I have money, then I would surely do the test.]*

Overall, health care professionals do not have a clear idea of the cost of the services they provide. *[I: What is the cost of a blood test for haemoglobin, HIV, syphilis and hepatitis B? Can you tell us the exact cost? HW6: No, I have no idea.]*

### Community-level

#### Taboo around sexual health

Most pregnant women, husbands and mothers-in-law were reluctant to talk about STDs. They laughed nervously, cut the discussion short or refused to answer questions. Also, the topics discussed were one of the main reasons why people refused to participate in the interview. This attitude of people not being ready to open about the subject was also found in the interviews with health professionals.

Many interviewees pointed out that women may feel embarrassed to talk about sexual health, even with doctors. *[HW1: Even patients who come here cannot talk openly about their reproductive health problems. Sometimes, they come to us and secretly tell us their problems.]* Especially if the doctor is a male. *[PW2: If there is a female doctor, then we can talk openly about our problems. Even if there is no female doctor, then we should not feel shy with the male doctor because if we feel shy then we cannot survive. But it would be better if there is a female doctor instead of a male doctor.]* This was often a reason given by participants for the need for the husband or someone to accompany the woman. *[H7: If there is a male doctor, then it can be difficult for the woman. The husband should go to make the situation easier.]*

#### Discriminatory attitudes and behaviours

Discriminatory behaviours regarding hepatitis B and syphilis were difficult to capture, as people were generally unaware of these particular diseases. However, the interviews revealed strong community stigma and peer pressure on HIV-positive people. *[H9: Some might feel the fear of embarrassment in society if the result comes positive because there is a practice of showing negative attitudes and discriminatory behaviour towards infected people in our society. Pregnant women might not agree to do the test due to the fear of being mistreated if they get diagnosed with STDs.]* HIV-positive people are often discriminated against and even excluded from society. *[HW2: The stigma here is that if you have contracted HIV, your life is over. How will I show my face? How am I going to get around?]* Thus, several participants emphasised the importance of awareness-raising programmes at the community level. *[PM1: Providing drugs is not enough. The main thing is to raise awareness. If people are aware, they can take care of themselves during pregnancy.]*

### Institutional level

#### Service delivery

The service delivery building block (30) focuses on providing equitable access to essential health services that meet the needs of the population by guaranteeing the availability of resources and the quality of the services provided.

#### Availability of services

The availability of blood tests depends on health facilities. Blood tests are done at the first ANC visit, usually after the third month of pregnancy. STDs are tested during pregnancy with other tests such as blood group or haemoglobin, using the same blood sample. The content of the blood tests depends on the healthcare facility. *[HW3: The test for HIV was considered important but not the ones for syphilis and hepatitis B. With time, the demand for other tests also gradually increased. Now, the ANC check-up package includes haematology, biochemistry, and serology tests.]*

Participants reported long waiting times in some cases, but this did not seem to discourage them from accessing ANC services. *[I: Sometimes it takes 2 or 3 hours to meet the doctor and sometimes even a whole day without eating any food.]* Waiting times for ANC visits, blood tests and results vary from one health facility to another. According to interviews, waiting times for the test can range from zero in some health posts to several hours in public hospitals for ANC visits, and from one hour in health posts or some provincial hospitals to several days in the PMWH to receive blood test results. Distance to health facilities did not appear to be an issue for blood testing. Participants interviewed in the PMWH sometimes reported coming from far away to access that particular hospital. *[H9: I came with my wife on a motorbike. If I am alone, it takes about 1.5 hours. But I had to ride slowly with her as she is pregnant and sick, so it took around 2.5 hours.]*

Many interviewees referred, directly or indirectly, to the referral of patients from lower- to higher-level facilities. One example that emerged repeatedly in the interviews was the referral of patients from health posts that do not have laboratory or ultrasound facilities, or in case of complications during pregnancy, to public hospitals. *[HW4: We don’t have ultrasound facilities. If an anomaly is detected in time, we are obliged to refer them elsewhere. They are referred to other services depending on their needs.]* Patients are also referred from one department to another within the same hospital. Several problems with the referral system emerged from the interviews. It sometimes makes it difficult for people to access care, creates confusion and leads to loss of follow-up during pregnancy. *[HW4: They find it complicated when we refer them to the hospital.]* This also sometimes leads to the saturation of the largest public hospitals. *[HW4: Everyone knows PMWH, but it is crowded most of the time.]* This referral system is sometimes used as an excuse for not taking care of HIV-positive patients. *[HW2: They would not admit an HIV-positive patient. They would still refer him to another health facility.]*

#### Quality of care

Overall, participants reported a high level of overcrowding in public hospitals that influence the quality of the services provided. *[PW12: There is so much crowd that patients enter the room for their turn before the previous one has completed the check-up. We can’t ask for anything and doctors don’t have time to explain things.]*

Even when blood tests are carried out, the quality of services is not always good. Some participants reported errors, receiving other people’s results or unreadable reports. *[PW11: When I had a miscarriage, I did a blood test in a polyclinic nearby and my blood group was different. Since then, I don’t want to do any tests there again because the report might have been exchanged with someone else.]* Others complained that the results of the blood tests were not explained to them by health workers. *[PW12: I came back after three days to get the report, but they did not tell me anything about it. I asked them whether I should go to ask about the report. Nobody told me anything about that, so I did not go anywhere.]*

Most of the participants said that they had not been counselled about blood tests and STDs. *[PW2: Health workers didn’t tell me anything. They just took the blood sample. It was very crowded.]* When counselling is provided, it is for HIV, but not for hepatitis B or syphilis. *[HW6: The counselling is about the prevention of HIV transmission from mother to child. I do not need to tell them about hepatitis B and syphilis.]* According to health workers, this depends on the health facility. For example, Tribhuvan University Teaching Hospital has a nurse who specialises in counselling, while health workers at other health facilities involved in ANC visits reported not doing counselling at all. Health workers justified not counselling pregnant women by the lack of time or space to do so, but also by the lack of interest or capacity to understand from the patients. *[HW7: In most health facilities, there is limited time for counselling. We cannot give half an hour or an hour to a patient when we have to deal with 100 or 150 patients. Sometimes, when people hear about HIV, they become reluctant, so we have to explain it to them. For hepatitis B, if patients understand and are willing to listen, then we explain to them too. Counselling is not done uniformly between patients, sometimes the explanations are done correctly but sometimes they are not.]* Some health workers also felt that it was easier not to mention STDs, as this would lead women to refuse blood tests. *[HW1: The easiest way to counsel them about blood screening is to tell them that the test should be done to see if there is a problem in the blood. If we explain each test to them, they may not be able to understand it. Also, mentioning STDs may make them angry because they may feel insulted. They don’t readily accept such tests.]* However, most health workers who report not counselling patients also say that they are happy to answer women’s specific questions about blood tests. *[HW4: We tell them that we will do a blood test. If they ask, we tell them that we will do these and these tests and tell them everything, but they never ask this kind of question.]*

Two health workers in the rural area cited language as a barrier to providing services to individuals, primarily counselling. *[HW1: At first, I was confronted with the problem of the Awadhi language but little by little, I managed and adapted to it. I’m still struggling to make sentences, but I can speak well enough to communicate.]*

### Health workforce

The building block (30) that considers health workforce addresses the availability, distribution, and capacity of healthcare workers.

#### Training and shortage of health workers

Most interviews revealed a shortage of healthcare workers, especially skilled workers. Many participants pointed to the large number of patients that need to be cared for by a limited number of health workers. *[H1: Thousands of people go to hospitals every day, so they are quite quick and give you information on the subject very quickly.]* This shortage results in a lack of time to provide health services and threatens their quality. *[PM4: Most of the ANC rooms are really busy. There will be women inside the room and the process of ANC is less than 5 minutes.]* Participants also complained about the lack of follow-up by doctors. *[PW9: You want to show your report to the same doctor, but it doesn’t happen that way. Even if it’s the same day, the doctor changes from time to time.]*

Training of health workers is usually provided by the health facilities themselves or by the municipality. NGOs are also sometimes responsible for training health workers on specific topics. Some health workers reported that they had not received training for decades. *[HW6: After 2015, we did not receive any training. I had a 7-day training on the prevention of mother-to-child transmission of HIV and a 10-day training on breastfeeding, but it was about 8-9 years ago.]* Almost all complained about the lack of regular training. *[HW4: Nowadays, the municipality provides the training, but it is not enough to improve the skills like it used to be before even if routine training is provided to health workers.]*

#### Attitudes of health workers

Regarding the attitude of the health workers, most pregnant women and their relatives said that the health workers had the right attitude, often described as kind. *[PW7: They are always cheerful and behave decently with me.]* Some participants reported inappropriate behaviour by health workers, such as rudeness or hostility toward pregnant women. *[PW12: Sometimes, when we ask them about anything, they scold us so badly for not knowing it. Once, I asked them about where I can get the blood test reports from. They shouted at me so badly asking why I don’t read myself and whether I have eyes or not. I felt very bad. Some are good but some of them are very aggressive.]* Also, some participants complained about the lack of listening by health workers. *[I: Have you ever experienced someone who behaved badly with you? PW5: Yes, sometimes they do not listen to you at all.]* With respect to STDs, health professionals were generally reluctant to talk about discriminatory behaviour in their profession. However, some decision-makers and health workers revealed that this type of behaviour is quite common. *[HW2: Health workers still act rudely sometimes. Sometimes clients even refuse to enter health facilities. Even HIV-positive people with health insurance are reluctant to come because the health staff do not treat them properly and send them from one place to another.]* They also indicated that the situation had improved. All health workers reported that they strive to maintain confidentiality, but face difficulties in practice, mainly due to lack of space.

The interviews revealed that, overall, people trust health workers. They often indicated that they give them good suggestions. *[ML3: Health workers in the health post will give her good suggestions. If we take suggestions, we can get good treatment and the right medicines.]* Participants said they were willing to listen to them and do what they recommended because they had better knowledge. *[ML2: We are uneducated, and we do not understand anything. If it affects the health of a pregnant woman as well as the child then, we will follow the doctor’s suggestion. We will try to follow whatever they tell us.]* However, some people suggested that people sometimes did not trust them and preferred to seek confirmation at different health facilities. *[PW2: The doctor said that there was no heartbeat. I did not believe it. The next day, I went to Bhairahawa for an ultrasound. They said the same thing there too.]*

### Health information

The health information building block (30) involves the collection, analysis and dissemination of data and information relating to health in order to facilitate decision-making and monitor the health of populations.

In Nepal, women’s maternity data is only reported after three months of pregnancy. Any event or illness occurring before the third month of pregnancy is not recorded in the Integrated Health Management Information System (IHMIS). The recording of STD data is done manually by health workers in all health facilities. While HIV data seems to be systematically recorded, this is not the case for syphilis and hepatitis B. According to some health workers, the lack of available information on syphilis and hepatitis B is due to the lack of available test kits and the lack of staff and time to record the data. *[HW7: In all facilities, data is reported manually. Patient loads are high, outpatient departments are busy with sometimes 200 to 300 patients coming and manpower is limited. We try to record data as much as possible, but we do not write the data both on the patients’ cards and in our data.]*

### Medical products

The building block (30) that considers access to essential medicines addresses the availability and appropriate use of medicines, vaccines, and medical equipment, with a focus on quality, safety, and affordability.

Interviews revealed a general lack of resources. *[PW9: They told me there was no heartbeat. They advised me to get admitted but due to a lack of beds at the emergency service, they could not admit me.]* While HIV testing kits are generally available in health facilities, syphilis and hepatitis B testing kits are often not available due to shortages. *[HW3: HIV test kits are usually available but not kits to test hepatitis B and syphilis. They are not funded by any program, and we cannot afford them. HIV kits are available through external funding.]* Laboratory services are often not available at health posts. *[PM4: We have ANC all over Nepal but screening for STDs is available only wherever there are laboratory facilities. That will be less than 200 government health facilities, hospitals, and a few Primary Health Care centres. The availability of ANC is very good in Nepal but when you go deep into the service and look at the quality and the components. Then what we call availability becomes poor.]* Many health workers also complained about the lack of appropriate counselling spaces. *[PM2: There are counselling rooms in the hospitals, but not in small health posts.]*

### Interpersonal level

#### Husband’s willingness to support

Overall, the husbands interviewed expressed a willingness to be involved in their wives’ pregnancies and often attested to supporting their wives with daily tasks. *[I: Do you support your wife during her pregnancy? H4: Yes, I support her 100%. I do not let her carry heavy objects or wash clothes. I encourage her to rest properly and follow doctors’ advice.]* However, some husbands reported not having any information about their wife’s pregnancy other than that the baby is doing well. *[H2: She said that the baby is normal and healthy, so I didn’t ask her about anything else. We were more concerned about the baby, so I didn’t think it was important to ask about anything else.]* While some couples discuss the pregnancy openly, others do not discuss it at all with each other. *[H4: We spontaneously exchange health-related information, such as what we should do, whether we should go for a walk, etc.]*

In urban settings, some husbands reported accompanying their wives on ANC visits. However, in large urban hospitals, they are sometimes not allowed to enter the room for check-ups, even if they want to *[H6: Health workers should welcome both the husband and his wife. They should tell me about the problems of my wife and that I should not worry.]* Husbands often expressed their inability to be there as much as they would like to be for their wives because of their work and the pressure to bring money into the household. *[H6: Sometimes when I did not have time to go with her due to office obligations, she went alone. Otherwise, we always go together.]*

In the migratory context of Nepal, among the eighteen families interviewed, four husbands, living in Kapilvastu, were reported not living with their wives now or in the previous months but abroad or in another city. Communication difficulties within the couple are sometimes reinforced by the lack of proximity when the husband has to migrate. In this situation, communication sometimes takes place through the pregnant woman’s in-laws. *[PW1: I told my sister-in-law first, then she shared it with everyone else. I: Didn’t you share the news with others? PW1: No because I was feeling shy.]*

With respect to STDs in particular, most husbands expressed support for their wives if she had symptoms of an STD. *[H7: If women get infected with STDs, they need full support from their husband. Husbands should go with their wives as they could have problems like difficulty walking, and dizziness.]* Mainly if she needs their help in discussing her situation with health professionals. *[H3: Some women might feel shy. I will tell my wife that I will come with her. She should not be scared.]* However, several women and health professionals interviewed pointed out that in practice, some husbands are not as supportive and understanding. *[PW11: Although my husband is understanding, many pregnant women have husbands who do not understand or care about them.]* Most interviewees said that the husband should be the first to know about his wife’s sexual health problems and that husbands should also be tested as part of antenatal screening for STD. *[PW3: It is important and necessary that the husband take a blood test too. It will help to identify who is infected and how we can prevent it.]*

#### In-laws’ involvement

In joint households, which are very common in rural areas, the parents-in-law are involved in the pregnant woman’s health, mainly the mothers-in-law or sisters-in-law. Indeed, the topics of pregnancy, sexual health or STDs are more easily discussed with people of the same sex. *[I: Do you think she should tell her husband? PW1: Yes, and her mother-in-law or sister-in-law, but not her father-in-law.]* The involvement of in-laws may take the form of advice, pressure, or accompaniment during check-ups. *[HW3: If they are having repeated infections or lower abdominal pain during intercourse, they can’t share such things in front of their mother-in-law. The mother-in-law may try to normalise the problems and demoralise the patients by not giving importance to their problems.]* In contrast, some mothers-in-law support their daughters-in-law. *[ML1: It is normal to help my daughter-in-law like this. She left her father and mother and came to live with us as if we were her parents. We must help her and understand her pain.]* Pregnant women generally need the permission of their in-laws before making decisions about their health. *[I: If the doctor says that they need to take your blood for STD tests, what would you do? PW3: I will say that I will decide after consulting with my family. I will ask my relatives if they want it or not. I will do as per their wish. I can’t just do whatever I want.]* In the context of migration, the husband often entrusts his wife to his family. *[H1: Our family lives together, so even if I can’t help and support her, my family supports her when I’m not there.]*

#### Neighbours’ involvement

In rural settings, neighbours are also an important source of information about health in general, the availability of health services and pregnancy in particular. *[PW5: My neighbour told me to go for a check-up if I had any health problem.]* In the case of STDs, participants shared that they learned a person’s STD status from neighbours. *[I: Have you ever heard of anyone infected with HIV, syphilis, or hepatitis B? PW12: My neighbours said that someone has such a disease so we should not visit him, otherwise we would get infected too.]* They play an important role in spreading rumours about the health of individuals and stigmatising behaviour. *[H2: There was a rumour in the village that he has HIV. We cannot go to visit him after knowing this about him.]* This type of behaviour seemed to be more present in rural areas and was not reported by participants in the Kathmandu Valley. *[H1: In urban areas like Pokhara, everyone is busy with their own work, why should I interfere in other people’s life?]*

### Individual-level

#### Health negligence

Some participants reported that they follow health workers’ recommendations carefully, go for regular check-ups, and comply with treatments. *[PW1: I usually go for ultrasounds when they call me. I’ve never missed an appointment.]* On the contrary, some participants, through the description of certain situations or more directly the health workers interviewed, revealed the negligence of individuals in health-related matters. *[PW4: The doctor told me to visit again but I did not.]* In the case of family members and given their involvement in the pregnancies described above in rural settings, their neglect has direct consequences since women’s health and consultations depend on them. *[PW2: I asked my in-laws to go for another check-up because it might be a problem with medication, but they did not take me for the check-up and then I stopped the medicines.]*

Another established norm in Nepal that emerged from the interviews is that if there are no significant symptoms, people believe that there is no disease or that it will heal on its own or with home remedies. *[HW3: They first follow all the possible home remedies they can. If nothing works, then only they come to the hospital for help.]* Thus, people only go to the hospital if the illness is severe, and the symptoms are important. *[HW2: Here we have a scenario where patients only go to the hospital when their condition is really serious. For example, with STDs, if patients have severe back pain or abnormal discharge or something serious, then only they feel the need to go to the hospital.]* People also tend to hide their health problems in general and especially when it comes to sexual health. *[PW11: Usually, people keep their disease secret, mostly in the village areas. It is similar in the case of miscarriage. For example, I did not reveal mine.]*

#### Knowledge about blood tests and STDs

When asked about their knowledge of ANC check-ups and blood tests, most participants were aware of their importance, although they did not know what was diagnosed by blood tests. When asked about their knowledge of STDs, most participants said they had heard of HIV and had some vague knowledge of the mode of transmission or symptoms. A limited number of educated participants have a good knowledge of the subject, mostly in urban settings. *[H9: HIV is a disease which gets transmitted through blood and sexual activities. It can get transmitted through blood by the use of needles, razors and sharp instruments if they are not sterilised.]* However, there is generally some confusion and misconception. *[H2: HIV can be transmitted by sitting and talking together. It can also be transmitted through sex or by living and sleeping together. If someone has this disease, we should not sit or live with him. We should keep our distance from such people.]* Participants reported hearing about it in the media, primarily on the radio, or during their studies. Most participants had never heard of syphilis and hepatitis B and had very low knowledge and awareness of their symptoms and mode of transmission.

#### Acceptance of blood tests

Consent for blood screening during pregnancy is given verbally by pregnant women. Health workers reported that blood tests were generally well accepted. This was confirmed by the pregnant women, husbands and mothers-in-law interviewed. The main reason given for the acceptance of blood tests was their importance to the health of the child and the mother. *[ML2: I would strongly advise the pregnant women to take a blood test timely as it helps to know if the health of the baby and mother is going well.]* Most of the people interviewed said they would accept the blood test, regardless of its purpose. This view was not shared by health professionals, who felt that most people would refuse it if it was explicitly explained to them that it was to test for STDs. *[HW2: If you organise a campaign to test blood for dengue, diabetes and malaria, they will readily agree to take the blood test. However, they will hardly agree to the same blood test for STDs. They would rather run away or scold you. They will certainly not agree to take the blood test.]* Potential reasons for reluctance to accept blood tests that emerged from the interviews were financial constraints, but also the fact that people do not consider them necessary because they are healthy or, conversely, they fear the results. *[H1: There are many reasons for not agreeing to have a blood test: for fear of what people might say, for fear of the disease, and in some cases, some people may feel that it is not necessary.]*

#### Deciding which health facility to visit

People reported preferring to receive all their care in a single establishment. [*HW4: Patients feel that they would feel more comfortable if all services were provided in this health facility, including ultrasound, instead of having to go to hospitals.*] However, the decision to change health facilities during pregnancy sometimes comes from the individuals themselves. In rural area, people sometimes come to the health post with reports of test results from private or public hospitals to confirm interpretations. This shows people’s trust in local health structures. *[H9: We took with us the report of the blood test we did at Binayak Hospital two or three days ago at the health post. Based on that report, they gave us some suggestions.]* In the Kathmandu Valley, people sometimes choose health facilities where to access ANC services, knowing that they will eventually be referred by health professionals. *[PW9: We thought that even if I go to a private hospital, they will refer me to this place if any complication arises, so why not come here directly.]* Participants also indicated that they went to larger public facilities, believing that the services provided would be of better quality, or to private clinics to avoid the crowds in public hospitals. *[HW6: People from elsewhere think that they have to come to this hospital to deliver properly and avoid complications during operations. They may think that there are few or fewer qualified doctors in their area.]* In the latter case, people usually come for a few ANC visits but cannot afford to do them all. *[HW7: Per visit, it costs 500 rupees, so they do one visit here and then we lose the follow-up. They think something like let’s pay for once to deviate from the government hospital line.]*

### Differences and similarities in perceptions between respondents in Kapilvastu (rural plains) and Kathmandu (urban)

Rural and urban settings differed in terms of respondents’ perceptions and the barriers and facilitators to antenatal screening identified. While, on the whole, the lack of resources in the health system emerged as one of the main barriers to antenatal screening in both urban and rural areas, the lack of STD screening services was reported more by participants in Kapilvastu, mainly at health posts. Similarly, referral of patients from health posts to public hospitals with laboratories was more common in rural areas. All health workers said that they tried to maintain confidentiality, but health workers working in Kathmandu said that they encountered difficulties in doing so, mainly due to lack of space. In Kathmandu, many health workers also complained about the lack of appropriate counselling facilities. This issue did not come up in the Kapilvastu interviews. In urban areas, participants highlighted the large number of patients who have to be cared for by a limited number of health workers, mainly in large urban hospitals, and participants complained about waiting times and husbands not being allowed to enter the room with their wives for examinations, even if they wanted to.

While husbands in both rural and urban areas expressed their inability to be as present as they would like for their wives, the reasons cited differed between the two settings. In Kathmandu, the main reason was work. In rural areas, husbands often lived abroad or in another town. In rural areas, joint households are more common than in Kathmandu, which makes the involvement of in-laws in the pregnant woman’s health more important there. Similarly, the involvement of neighbours as a source of information, the spreading of rumours about people’s health and stigmatising behaviour were not reported in the Kathmandu valley, whereas they were by several participants in Kapilvastu. While the attitude of people not being ready to open up on the subject of STDs was found in both rural and urban areas, this finding was more exacerbated among participants in Kapilvastu. Some health workers in Kapilvastu were not even prepared to say the words “sexually transmitted diseases”.

### Differences and similarities in perceptions between different household members

In interviews with the members of the same households some interesting similarities and differences between household members’ perceptions emerged. Husbands and mothers-in-law were generally less reluctant to talk about pregnancy and their knowledge of STDs than pregnant women. Pregnant women were particularly reluctant to talk about the subject, even if the interviewer was of the same sex. In the joint households interviewed in Kapilvastu, most of the pregnant women were embarrassed to talk about their mother-in-law’s involvement in their pregnancy, even if she was not in the room, whereas the mothers-in-law were not embarrassed to talk about their daughter-in-law.

Husbands expressed support for their wives if they had STD symptoms, while pregnant women and mothers-in-law tended to perceive husbands’ support as weak in some cases, as husbands were not as understanding as they might say. The main reason given by husbands for accepting blood tests was their importance for the child’s health, while the health of both mother and child was mentioned by mothers-in-law and pregnant women. Husbands and pregnant women agreed that they wanted the husbands to be involved in their wives’ pregnancies but tended to differ in their perceptions of in-laws’ involvement. Some pregnant women complained about the involvement of their mothers-in-law and sisters-in-law in their pregnancy, particularly in Kapilvastu, while husbands and mothers-in-law were quite satisfied about it.

## Discussion

This qualitative study showed that integrated antenatal screening for HIV, syphilis and hepatitis B in Nepal involved many stakeholders and was influenced by factors at different levels of the socioecological model and across the different WHO building blocks. Barriers to antenatal screening were mainly on the supply side, including a general lack of resources, a shortage of healthcare workers and a lack of training. However, husbands and in-laws also played an important role in the acceptance of screening by pregnant women in the Nepalese context, mostly in the rural setting.

Integrated screening for HIV, syphilis and hepatitis B in Nepal is part of the National HIV Testing and Treatment Guidelines and it is mandated to be offered free of charge to all pregnant women during their ANC visits (11). However, there is a lack of information on the coverage of screening for syphilis and hepatitis B, which is not included in HMIS or national surveys, making the investigation very difficult. The implementation of HIV screening is limited and uneven across regions. In Gandaki Province, 25% of pregnant women received an HIV test during ANC compared to only 7% in Karnali Province (34). This is mainly due to competing priorities at local level with the social sector, including health, no longer taking priority over other directly visible investments such as road or building construction, which would encourage the re-election of decision-makers (35,36). Political instability, which makes it difficult to implement long-term decisions (37), and a general lack of resources (15) are other main factors. In provincial hospitals, this lack of resources is mainly reflected in a lack of space, which prevents counselling in private, and a shortage of qualified healthcare workers. These results were found by other studies conducted in Jumla (38) and southern plains in Nepal (39). Insufficient healthcare workers translates into a lack of time to provide health services, which threatens service quality and increases women’s waiting times. In lower-level healthcare facilities, such as health posts, a lack of laboratory services or screening equipment, especially for syphilis and hepatitis B is a major problem. Overall, health workers suffer from a lack of appropriate training on STDs which is important to reduce stigmatising behaviour (40,41).

Local authorities play an important role in the implementation of integrated antenatal screening. Despite an overall lack of clarity in roles and responsibilities between different levels of government since the federalisation in 2017 (42), municipality-level health coordinators and the elected officials in-charge play a key role in the implementation of health protocols and allocation of funding. Yet the elected officials often lack technical training or experience to inform their decision-making (36) and integrated antenatal screening for STDs was often not a priority for decision-makers (15,36). Meanwhile, the federalisation increased the number of layers for decision making and the high administration burden increase the time required to introduce changes (36). As in other areas such as education, the move to federalism can also create tensions over the division of authority between federal and local government, reducing the effectiveness of decision-making (36,43).

Our study reveals the importance of the referral system in Nepal for ANC and blood tests during pregnancy and especially in rural areas where access to screening services is limited (44). Referral systems are meant to ensure that patients receive appropriate and timely health services, but it is sometimes high levels of referral are responsible for hospital services becoming clogged with ‘normal’ cases that are not in need of specialised care. The lack of screening facilities in lower-level facilities plays a large role in the process. Despite this, we found that individuals have high trust in local health facilities which suggests that there is an opportunity to enhance screening and counselling at health posts to reduce the need for referral. Massive investment in laboratories in public health facilities at the lower level is needed to implement triple screening at an acceptable level.

HIV screening during pregnancy falls under the Prevention of Mother-to-Child Transmission program (45), implemented by the Ministry of Health and Population of Nepal with support from various international organisations, including UNICEF, the World Health Organization (WHO), and the United States Agency for International Development (USAID). This program is independent from the ANC program. Among the three diseases we consider, HIV received a larger coverage in terms of screening resources, awareness campaigns and media coverage. HIV screening is of high interest worldwide and Nepal has received substantial financial aid in the last decade from the international community (46). However, this is not the case for syphilis and hepatitis B despite higher prevalence of these diseases compared to HIV in the country (6). A systemic lack of resources for antenatal screening of syphilis and hepatitis B and the fact that data are not systematically reported for these diseases (47) means that these diseases and their transmission from mother to child remain woefully neglected. The fact that facilities are less able to provide hepatitis B and syphilis screening services than HIV or ANC services indicates that these services have not been fully integrated (15). If rapid tests exist for HIV and are used in Nepal, a similar investment in the provision of rapid tests for hepatitis B and syphilis could facilitate antenatal screening and increase screening uptake (48).

The priority given to HIV over other diseases is reflected in people’s knowledge. We found relatively good knowledge about HIV compared to syphilis and hepatitis B. However, the lack of knowledge on the transmission of HIV and misconception about the risks of transmission lead to high stigmatisation of HIV-positive people within families and in the community. Overall, we found a large taboo surrounding sexual health and STDs, as found by other studies conducted in Nepal (49,50). These taboos lead to discriminatory behaviour. Generally, people delay care seeking in Nepal but when it comes to STDs, individuals further delay screening and care due to shame and fear of judgement. Counselling for HIV has been shown to be an effective strategy to reduce stigma and increase community knowledge (51). However, in Nepal, counselling seems to be case-by-case with women usually not informed on the purpose of the blood tests they are having. Our findings underscore the importance of awareness programs at the community-level which might help make up for the lack of resources for individual counselling. To reduce stigma, community awareness campaigns will need to be accompanied by the promotion of a culture of transparency by healthcare professionals towards patients regarding the objectives of screening and the importance of sharing screening results. The more health professionals associate themselves with stigmatisation by not mentioning diseases to patients, the slower the resolution of stigmatisation may be.

Financial barriers prevent access to, and acceptance of, screening. Financial barriers to antenatal screening were identified in other Asian studies (52,53). People move between facilities to save money even when this implies long waiting times in public hospitals. Switching between providers can break continuity of care and increase crowding in governmental hospitals. Individuals’ choice of health facility can be driven by a wide disparity in prices and quality between public and private facilities (54,55) and is often influenced by neighbours or family members (56,57).

We found that the husband’s lack of involvement or support for antenatal screening or more generally during pregnancy is often due to a lack of time, the result of pressure to bring money home, rather than an intention to withdraw support. Husbands’ lack of engagement is reinforced by migration, especially in rural areas (19). In Nepal, women usually move into their husband’s households after marriage (58,59). Her decision-making power in a household that is not her own is often weak (60,61). This trend is changing in recent years, mostly in urban contexts with people no longer living in joint families. However, social position within the household remains an important determinant of decision-making capacity in Nepal (62,63). The migration of husbands often complicates the communication between spouses and increases the involvement of in-laws in women’s pregnancies. We found that women in joint households generally need the approval of their in-laws before making decisions about their health, which has also been found by others (64,65). This again suggests the importance of community-level interventions to raise awareness about the importance of triple screening in pregnancy as well as general understanding about ANC.

Our study has several limitations. Firstly, in Kathmandu, participants were recruited through the PMWH, which meant that all the women interviewed had already attended at least one ANC visit and we were not able to access women who had not received ANC. Secondly, given the sensitivity of the subject, people were sometimes reluctant to answer questions about STDs. When people said they did not know about HIV or how it was transmitted, it was almost impossible to determine whether this was a real lack of knowledge or shyness about discussing the subject. Although we planned to interview husbands, mothers-in-law, and pregnant women from the same households, this was only possible in a subset of households in Kapilvastu and in none of the cases in Kathmandu. This limited our capacity to triangulate and compare perceptions of different household members within households.

Despite limitations, our study offers unique insights on factors influencing integrated antenatal screening for HIV, syphilis, and hepatitis B in Nepal. Improving integrated antenatal screening will require a multi-sectoral approach that involves strengthening healthcare systems, addressing stigma and discrimination, and increasing investment in syphilis and hepatitis B prevention programs. It will require greater engagement with communities through awareness programs and enhancement of the role of health posts, especially in big cities, to regulate the flow of patients in large public hospitals. Decongestion of the public health system might also be facilitated by regulation of prices for services in the private sector.

## Conclusion

Our study demonstrated that numerous interrelated factors affect integrated antenatal screening for HIV, syphilis, and hepatitis B in Nepal. Although supply side such as a lack of resources or a shortage of health workers for screening were important, we also found that husbands and in-laws play a key role in women’s screening decisions. Findings highlight the need for investment in the sector, increased training for health workers and awareness campaigns for men and women at the community-level.

## Data Availability

Fully anonymised transcripts are uploaded to the UCL research data repository. All transcript data will be provided on request.

## Acknowledgments

None.

## Supporting information

**S1 File. Vignette 1 - A pregnant friend heard about ANC visits**

