## Supplementary material for "Factors influencing the implementation of integrated screening for HIV, syphilis, and hepatitis B for pregnant women in Nepal: a qualitative study": Supplentary Material 1

### Vignette 1 - A pregnant friend heard about ANC visits

Imagine that a close friend named Sarita visits you at home for tea. You are alone in the room; the rest of your household is going about its business. She is about the same age as you, living in your neighbourhood and she is pregnant too. She has heard about ANC visits on the radio and wants to know your opinion.

Sarita asks you what you think about ANC visits. What would you answer?

Sarita asks you where she can go to have an antenatal check-up. What would you answer?

She asks you if she is going to have to pay anything. What would you answer?

How would you describe to her the process of an ANC appointment from the time the appointment is made to the time the woman leaves the health facility?

- How long will she wait and where?
- Should she go alone?
- What will health workers do?

*[If the interviewed woman refers to blood tests during the ANC visit directly, ask the following questions. If not, suggest that Sarita may need to have blood tests done during the visit for various things and then ask the following questions]*

- Will the tests be explained?
- What will be the tests for?
- How will the results be communicated?

How long will she wait for lab results?

Why do you think Sarita could be reluctant to attend the ANC visit?

Why do you think Sarita could refuse blood tests during the ANC visit?

- Can you tell me any reasons that Sarita would refuse a blood test if she knows it is to test for anaemia?
- Can you tell me any reasons that Sarita would refuse a blood test if she knows it is to test for HIV, Hepatitis B or syphilis?
- Can you tell me any reasons that Sarita would refuse a blood test if she knows it is to test for STD?

How would you encourage her to go to an ANC visit?

What would you say to her if she is afraid of getting a blood test?
